# Estimating the impact of transmitted and non-transmitted psychiatric and neurodevelopmental polygenic scores on youth emotional problems

**DOI:** 10.1101/2023.06.26.23291893

**Authors:** Amy Shakeshaft, Joanna Martin, Charlotte A. Dennison, Lucy Riglin, Cathryn M. Lewis, Michael C. O’Donovan, Anita Thapar

## Abstract

Anxiety and depression (emotional disorders) are familial and heritable, especially when onset is early. However, other cross-generational studies suggest transmission of youth emotional problems is explained by mainly environmental risks. We set out to test the contribution of parental non-transmitted genetic liability, as indexed by psychiatric/neurodevelopmental common polygenic liability, to youth emotional problems using a UK population-based cohort: the Millennium Cohort Study. European (N=6,328) and South Asian (N=814) ancestries were included, as well as a subset with genomic data from both parents (European: N=2,809; South Asian: N=254). We examined the association of transmitted (PGS_T_) and non-transmitted polygenic scores (PGS_NT_) for anxiety, depression, bipolar disorder and neurodevelopmental disorders (attention-deficit/hyperactivity disorder [ADHD], autism spectrum disorder [ASD], schizophrenia) with youth emotional disorder and symptom scores, measured using the parent-and self-reported Strengths and Difficulties Questionnaire emotional subscale at 6 timepoints between ages 3-17 years. In the European sample, PGS_T_ for anxiety and depression, but not bipolar disorder, were associated with emotional disorder and symptom scores across all ages, except age 3, with strongest association in adolescence. ADHD and ASD PGS_T_ also showed association across ages 11-17 years. In the South Asian sample, evidence for associations between all PGS_T_ and outcome measures were weaker. There was weak evidence of association between PGS_NT_ for anxiety and depression and age 17 symptom scores in the South Asian sample, but not in the European sample for any outcome. Overall, PGS_T_ for depression, anxiety, ADHD and ASD contributed to youth emotional problems, with stronger associations in adolescence. There was limited support for non-transmitted genetic effects: these findings do not support the hypothesis that parental polygenic psychiatric/neurodevelopmental liability confer risk to offspring emotional problems through non-transmitted rearing/nurture effects.

## Introduction

Depression and anxiety are the most common mental health disorders and a leading cause of global disability^1^. Outcomes are worse when these disorders first onset in childhood and adolescence^2, 3^. It is well-established that both major depressive disorder and anxiety disorders are familial and show modest heritability^3, 4^. However, these disorders are highly heterogenous and vary in their age at onset, among other features. When depression onsets early, it appears more strongly familial and heritable^5–7^. In contrast, other research highlights the important contribution of environmental risk factors to anxiety and early-onset depression. First, rates of emotional problems and disorders (anxiety/depression) have risen sharply in young people across different countries in recent years. This rise is most likely attributable to non-genetic factors, although the reasons behind the increased rate remain unknown^2^. Second, cross-generational genetic designs (e.g., adoption, in-vitro fertilization, children-of-twins) have further shown that transmission of anxiety, depression and more broadly defined emotional problems across generations is likely accounted for by both environmental factors and inherited contributions^8, 9^, or by environmental transmission entirely, especially in childhood^10, 11^.

One recently developed approach to studying cross-generational genetic transmission uses genomic data from complete parent-offspring trios. This method enables assessment of the effects of both transmitted alleles (i.e. inherited) and non-transmitted alleles (known as “genetic nurture”^12^). For example, genetic nurture appears to be an important contributor to educational attainment^13^ but less important for ADHD^14^. Such studies suggest that parental alleles contribute to educational attainment not only through inheritance but also through environmental mechanisms^13^. Few studies have yet used this approach to examine anxiety and depression. One study estimated the total parental indirect genetic effect of genotyped common variants on offspring emotional symptoms at age 8, finding no evidence of maternal or paternal genetic nurture effects for anxiety and depression symptoms in 3,801 trios in the Norwegian Mother, Father and Child cohort (MoBa)^15^. A larger sample from the same cohort (N=11,598 trios), however found evidence of parental non-transmitted genetic effects on depression symptoms in their offspring at age 8, but not on anxiety^16^. As the authors acknowledge however, depression symptoms are rare in prepubertal children.

When considered together, genetic studies show mixed findings. Some highlight a stronger genetic contribution when onset of emotional problems (at least depression) is early, whereas other designs observe strong environmental contributions to their cross-generational transmission. Convergent evidence, preferably across multiple study designs, is needed to better understand the etiology of emotional problems. Ideally, etiology needs to be examined at different ages, given that depression shows differences in its genetic architecture across development^2, 17^. Also, to date, research has focused on European populations and further studies are needed in diverse, non-European ancestry cohorts.

In this study, we tested two hypotheses using parent-offspring trios from an ancestrally diverse UK population-based cohort: the Millennium Cohort Study (MCS). First, we tested the hypothesis that <u>transmitted PGS</u> for anxiety, depression and bipolar disorder, contribute to the onset of emotional problems across childhood and adolescence, with greater contributions in adolescence. We also test transmitted PGS for neurodevelopmental disorders (attention-deficit/hyperactivity disorder (ADHD), autism spectrum disorder (ASD), and schizophrenia) as these PGS have been observed to contribute risk more strongly for early-onset depression^17–19^. Next, we tested the hypothesis that <u>non-transmitted parental PGS</u> for depression, anxiety, bipolar disorder and neurodevelopmental disorders are associated with emotional problems across childhood and adolescence (ages 3 to 17 years), especially in early childhood (i.e. evidence of genetic nurture).

## Methods and Materials

### Sample description

We used data from genotyped individuals within the Millennium Cohort Study (MCS; www.cls.ioe.ac.uk/mcs), a nationally representative UK longitudinal population-based cohort of children born between 2000–2001^20^. The sampling design ensured adequate representation across England, Wales, Scotland, and Northern Ireland, as well as enriched selection of families from ethnic minority backgrounds and from deprived areas of the UK. Ethical approval for MCS was gained from the Multi-Centre Research Ethics Committee. Parents gave written informed consent. We used data collected at seven time points (or “data sweeps”) from 2001–2018, starting when the cohort members (children) were aged 9 months and subsequently at ages 3, 5, 7, 11, 14 and 17 years. The initial recruited sample consisted of 18,818 cohort members from 18,552 families. Additional eligible families were recruited in 2003–2004, to a total of 19,519 cohort members from 19,244 families. DNA samples were collected from families when children were aged 14 years^21^ (from 9,259 cohort members); for details see Supplementary Text. Within this genotyped sample, data were available for 3,373 complete parent-offspring trios. MCS survey data can be accessed by bona fide researchers through the UK Data Service (https://doi.org/10.5255/UKDA-Series-2000031).

### Outcome measures

1. Emotional disorder Emotional problems were measured using the Strengths and Difficulties Questionnaire, emotional subscale (SDQ-E)^22^ by parent-report on their children at ages 3, 5, 7, 11, 14 and 17 years and adolescent self-report at age 17. This subscale consists of five items (“often complains of headaches”, “many worries”, “often unhappy, downhearted”, “nervous or clingy in new situations”, and “many fears, easily scared”), scored as 0 (not true), 1 (somewhat true), and 2 (certainly true), with the total score ranging from 0 to 10. The SDQ-E is a validated measure of anxiety disorders and major depressive disorder in children and adolescents^23^, and has been shown to have scalar invariance across ethnicities in the MCS cohort^24^. In line with previous research validating the SDQ-E against clinical diagnoses, we defined probable emotional disorder using the ‘very high’ cut-off^25^ for parent-and self-reported SDQ-E (≥7). Our primary outcome was a binary variable, ‘emotional disorder’, indicating the presence of a probable emotional disorder, based on exceeding this cut-point at any time, by either rater.
2. Emotional problems: total symptom scores Total symptom scores for the parent-rated and self-reported SDQ-E were computed. To test how robust our findings were across different measures, we also generated total symptom scores for the self-rated Short Mood and Feelings Questionnaire^26^ (SMFQ, 13 items) at age 14, which measures depression symptoms, and the Kessler Mental Health Scale^27^ (KMH, 6 items) at age 17, which measures psychological distress.

### Socio-economic status

Socio-economic status (SES) was defined using the modified Organization for Economic Co-operation and Development (OECD) equivalized income scores at the time of the child’s birth and standardized as Z-scores to aid interpretation.

### Genetic data

Details of genotyping, imputation, and quality control (QC), including relatedness checks are provided in the Supplementary Text. Principal components analysis (PCA) was used to identify super-populations and define relatively homogenous ancestry groups. All cohort members within the two largest population groups (European (EUR) and South Asian (SAS)) and their biological parents were included. A total of 7,142 cohort members passed all QC (EUR: N=6,328; SAS: N=814), of whom 3,063 also had genetic data from both biological parents available (EUR: 2,809; SAS: 254). A total of 3,976,009 autosomal SNPs passed QC. For each child with genetic data available from both biological parents (i.e. complete trios), non-transmitted parental alleles were derived in PLINK^28^, to create a ‘pseudo-control’ containing the non-transmitted parental alleles.

We calculated transmitted and non-transmitted PGS using independent large-scale genome-wide association studies (GWAS), for the following primary phenotypes: anxiety^29^, major depressive disorder (MDD)^30^, broad depression^31^, bipolar disorder^32^, and neurodevelopmental disorders [ADHD^33^, ASD^34^ and schizophrenia^35^]. PGS were calculated using PRS-CS-auto^36^ (see Supplementary Text). Parental PGS for the same phenotypes were also generated using the same method.

### Statistical analysis

All analyses were conducted in R^37^. We started by examining sample demographics and the frequency of a probable emotional disorder in all genotyped cohort members, trios only, and stratified by ancestry. Chi-squared tests were used to test for differences between groups for binary variables and Mann-Whitney tests for differences in mean emotional problem symptom scores. To test for associations between PGS and outcome measures we performed linear regression models for continuous outcomes (emotional problem symptom scores) and logistic regression models for binary outcomes (emotional disorder), and co-varied for 10 ancestry-specific principal components (PCs). Analyses between non-transmitted PGS and outcome measures were only performed in complete trios. Analyses of transmitted PGS with outcome measures were carried out in the whole genotyped cohort, since parental genetic information was not required.

We used the false discovery rate (FDR) to correct for multiple comparisons across transmitted and non-transmitted PGS for anxiety, depression and bipolar disorder. We also corrected for multiple comparisons across transmitted and non-transmitted PGS for neurodevelopmental disorders. A q-value of 0.05 was used to define the FDR threshold. All analyses were conducted separately for EUR and SAS samples.

### Sensitivity analyses

We tested for sex differences in the associations between transmitted and non-transmitted PGS with the binary outcome of emotional disorder by including a sex*PGS interaction term in logistic regression models. We also present models stratified by sex in the Supplementary Materials. Further, to test for mother/father-specific effects, which may have been missed when studying overall effects, we included mother, father and child PGS (each residualised against 10 ancestry-specific PCs) in logistic regression models of youth emotional disorder outcome. This method estimates the effect of each parental PGS on the outcome whilst controlling for the child and other parent’s PGS, thereby allowing to test for parent-specific genetic nurture, and has been shown to be equivalent to the non-transmitted PGS design in estimating direct/indirect genetic effects^38^.

To investigate the potential influence of non-random mating on the results, we examined correlations between mother and father PGS, between transmitted and non-transmitted PGS, as well as the degree of relatedness in trio parents (see Supplementary Text). Correlations were carried out using PGS residualised against 10 ancestry-specific PCs. We also tested for associations between transmitted PGS and offspring emotional disorder in the subset of cohort members who had data available for the complete trio (i.e. the subset used for non-transmitted PGS analysis), as well as testing for differences in socioeconomic status and sex distribution between the two subsets (trios vs. whole sample) using a chi-squared test for binary variables and a t-test/Mann-Whitney test for continuous variables.

## Results

### Sample demographics

Descriptive demographics including sex, ancestry (EUR or SAS) and SES, for the whole genotyped sample (Total N=7,142; EUR N=6,328; SAS N=814) and the complete parent-offspring trio subset (Total N=3,063; EUR N=2,809; SAS N=254) are displayed in **Table S1**. In the whole sample, 29% (N=1,383) of cohort members met the cut-point for having an emotional disorder at any time point. This proportion differed by ancestry (EUR: 28%, SAS: 40%, χ^2^(1) = 20.1, p=7.5×10^-6^). Females were more likely to have an emotional disorder, compared to males, in both ancestry groups (EUR: females = 37%, males = 19%, χ^2^(1) = 167.5, p=2.5×10^-38^; SAS: females = 47%, males = 30%, χ^2^(1) = 11.2, p=0.0008). There was an equal sex split (1:1) in both ancestry groups. Cohort members with data available for complete trios were less likely to have an emotional disorder compared to the whole sample (24% vs. 29%, χ^2^(1) = 21.8, p=3.0×10^-6^) and had higher SES (**Table S1**), which was consistent across both ancestry groups. Mean symptom scores for SDQ-E, SMFQ and KMH were also lower in the cohort members with data available for complete trios, at all ages and in both ancestries (**Figure 1**, p-values in **Table S1**). Details of missing data for covariate and outcome variables can be found in **Table S2**.

**Figure 1.**
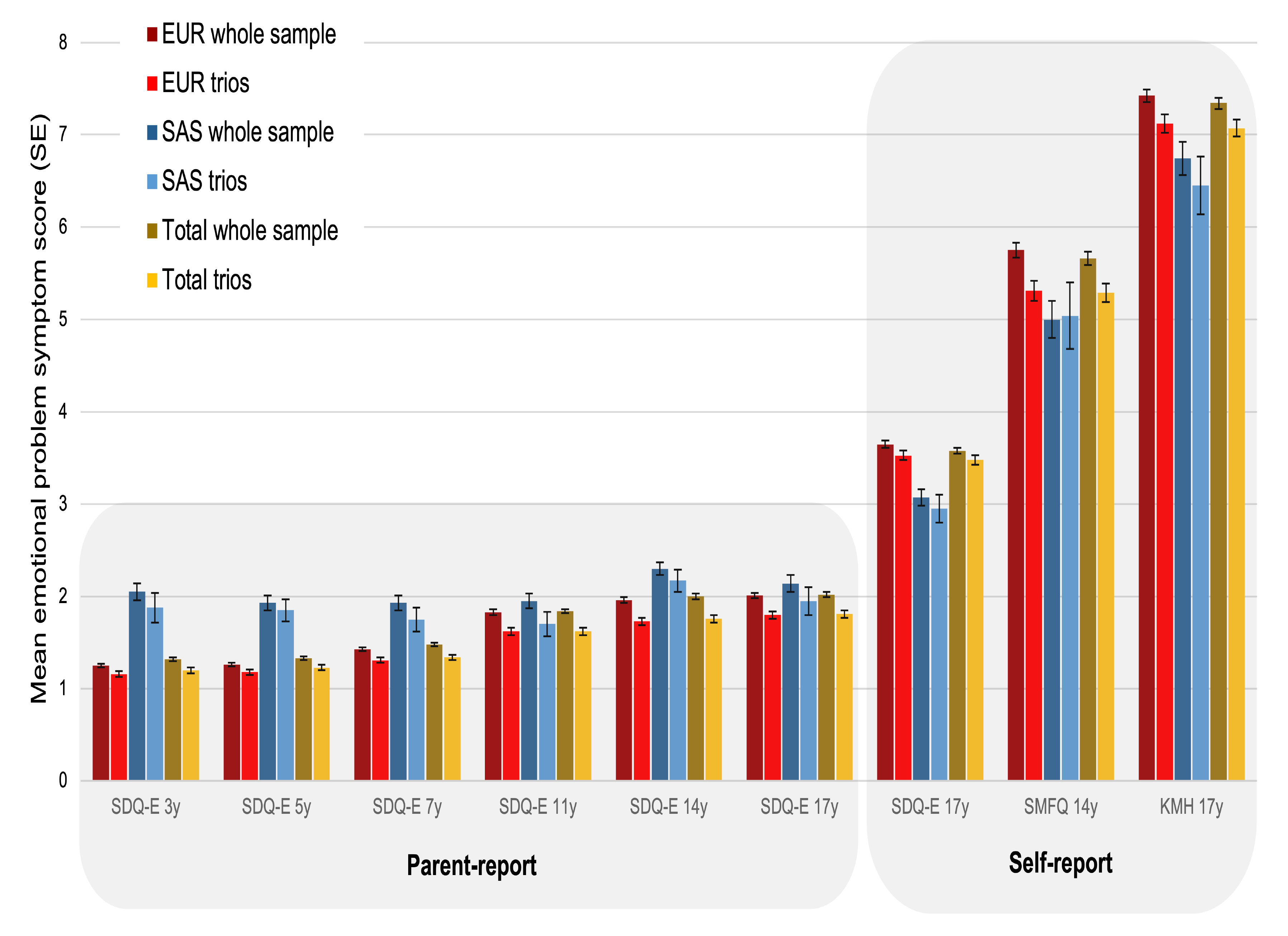
*Mean emotional problem symptom scores (±standard error [SE]) for parent-and self-reported mental health questionnaires in subsets of the MCS dataset. For results of tests for differences between trios vs. whole sample see **Table S1**. EUR: European ancestry; KMH: Kessler Mental Health Scale; SAS: South Asian ancestry; SDQ-E: Strengths and Difficulties Questionnaire emotional problems subscale; SE: standard error; SMFQ: Short Mood and Feelings Questionnaire.*

### Association of transmitted psychiatric/neurodevelopmental PGS with emotional disorder and symptom scores

#### Emotional/mood disorder PGS

In the European ancestry group we observed associations between transmitted PGS for anxiety, MDD and broad depression and youth emotional disorder (**Table 1**). These PGS were also associated with higher total symptom scores measured by SDQ-E, SMFQ and KMH at all ages, except at 3 years (**Figure 2a, Table S3**). Bipolar disorder PGS were not strongly associated with emotional disorder or symptom scores, except for an association with self-reported symptoms assessed using the KMH at age 17.

**Figure 2.**
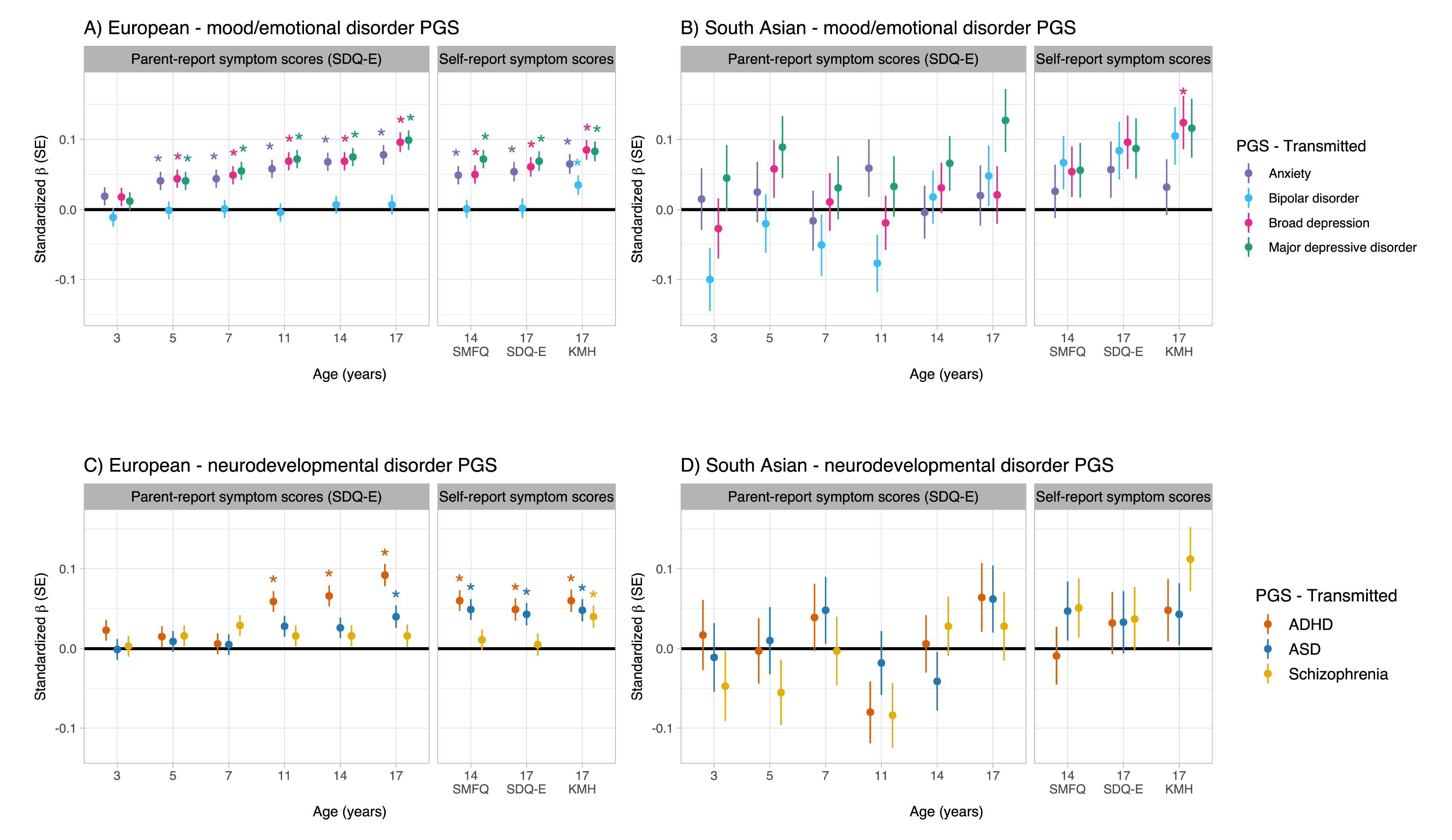
*Association between transmitted PGS for mood/emotional disorders and neurodevelopmental disorders with emotional problem symptom scores at different ages across childhood and adolescence (parent-and self-report), stratified by European (EUR) and South Asian (SAS) ancestry groups. ADHD: attention-deficit/hyperactivity disorder; ASD: autism spectrum disorder; PGS: polygenic score; SDQ-E: Strengths and Difficulties Questionnaire emotional problems subscale; SE: standard error; SMFQ: Short Mood and Feelings Questionnaire; KMH: Kessler Mental Health Scale. *FDR-corrected p<0.05.*

**Table 1.**
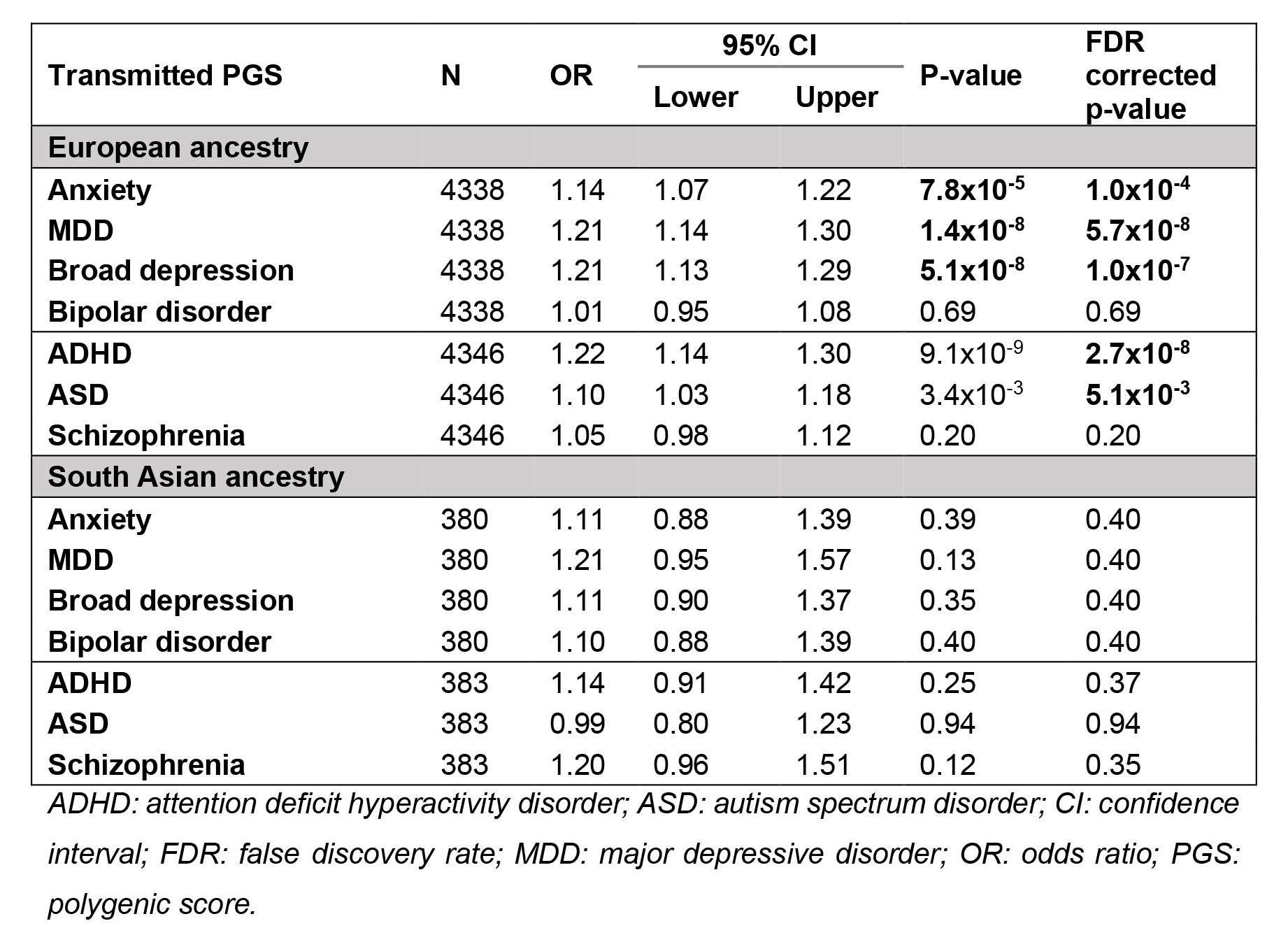
*Associations between transmitted psychiatric/neurodevelopmental PGS and youth emotional disorder in European and South Asian ancestries*.

In the South Asian ancestry group, there was not strong evidence of associations between transmitted PGS and emotional disorder (**Table 1**), with the exception of transmitted broad depression PGS and self-reported KMH at age 17 (**Figure 2b, Table S4**).

#### Neurodevelopmental disorder PGS

In the European ancestry group, transmitted ADHD and ASD PGS were associated with emotional disorder (**Table 1**). Transmitted ADHD PGS were also associated with parent-and self-reported symptom scores between ages 11-17 years. Transmitted ASD PGS were associated with self-reported symptom scores (ages 14-17 years) and parent-reported scores at age 17 (**Figure 2c, Table S3**). Schizophrenia PGS were associated with self-reported KMH at age 17 (**Figure 2c, Table S3**).

In the South Asian ancestry group, there was not strong evidence of association between any transmitted neurodevelopmental PGS with emotional disorder (**Table 1**), nor for symptom scores at any age (**Figure 2d, Table S4**).

### Association of non-transmitted psychiatric/neurodevelopmental PGS with emotional disorder and symptom scores

#### Emotional/mood disorder PGS

In the European ancestry group, there was not strong evidence of association with emotion disorder for any non-transmitted PGS (**Table 2**), nor for symptom scores (**Figure 3a, Table S5**).

**Figure 3.**
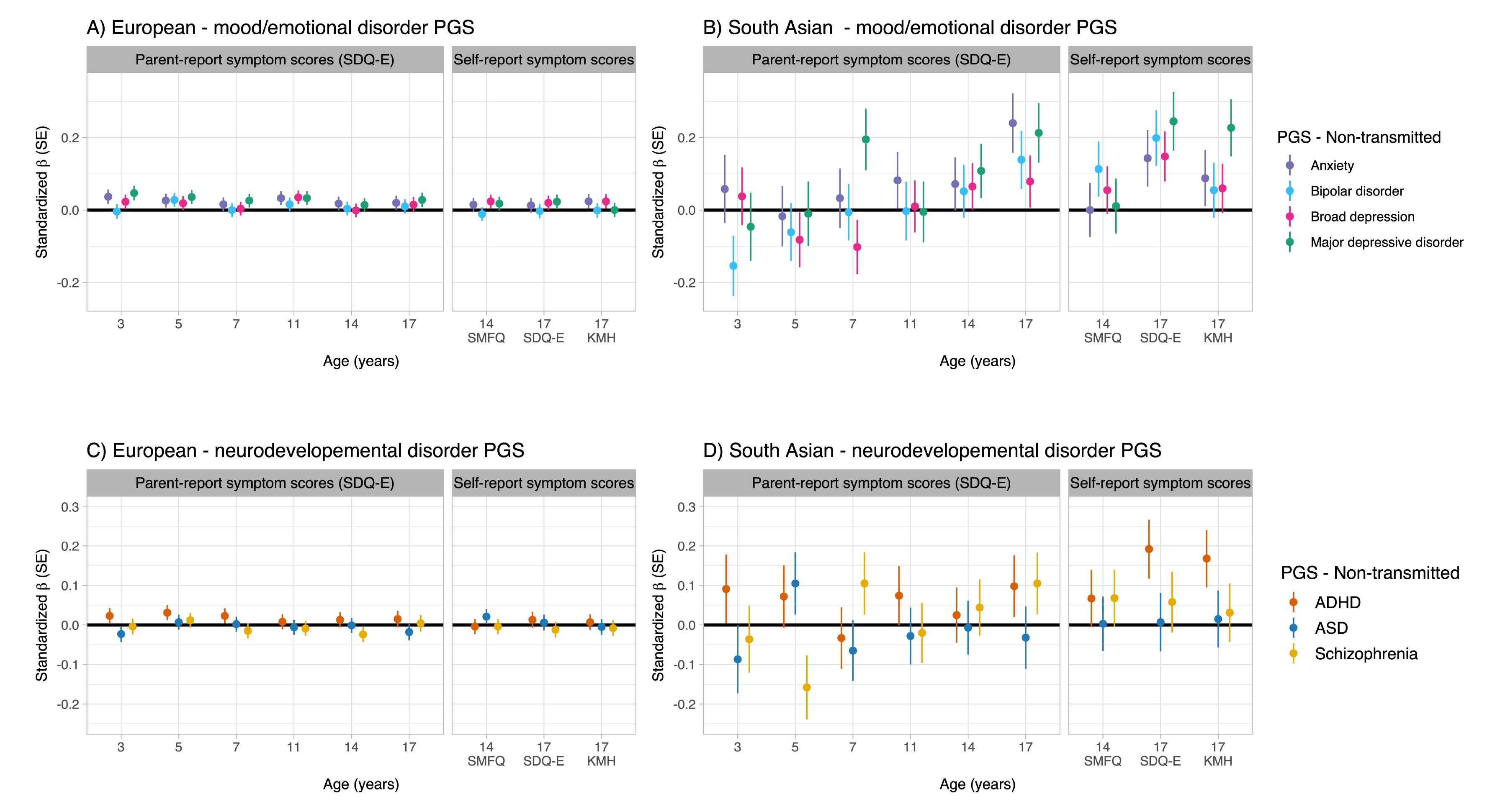
*Associations between non-transmitted PGS for mood/emotional and neurodevelopmental disorders with emotional problem scores at different ages across childhood and adolescence (parent and self-report), stratified by European (EUR) and South Asian (SAS) ancestry groups. ADHD: attention-deficit/hyperactivity disorder; ASD: autism spectrum disorder; PGS: polygenic score; SDQ-E: Strengths and Difficulties Questionnaire emotional problems subscale; SE: standard error; SMFQ: Short Mood and Feelings Questionnaire; KMH: Kessler Mental Health Scale. *FDR-corrected p<0.05*. Supplementary

**Table 2.** *Associations between non-transmitted psychiatric/neurodevelopmental PGS and youth emotional disorder in European and South Asian ancestries*.

| Non-transmitted PGS | N | OR | 95% CI |  | P-value | FDR corrected p-value |
| --- | --- | --- | --- | --- | --- | --- |
|  |  |  | Lower | Upper |  |  |
| European ancestry |  |  |  |  |  |  |
| Anxiety | 2129 | 1.00 | 0.91 | 1.11 | 0.98 | 0.98 |
| MDD | 2129 | 1.01 | 0.91 | 1.11 | 0.90 | 0.98 |
| Broad depression | 2129 | 1.03 | 0.93 | 1.14 | 0.55 | 0.98 |
| Bipolar disorder | 2129 | 0.94 | 0.85 | 1.04 | 0.21 | 0.85 |
| ADHD | 2133 | 1.04 | 0.94 | 1.14 | 0.50 | 0.74 |
| ASD | 2133 | 0.98 | 0.89 | 1.09 | 0.76 | 0.76 |
| Schizophrenia | 2133 | 0.94 | 0.85 | 1.04 | 0.22 | 0.66 |
| South Asian ancestry |  |  |  |  |  |  |
| Anxiety | 119 | 1.52 | 0.94 | 2.54 | 0.09 | 0.19 |
| MDD | 119 | 1.60 | 0.95 | 2.78 | 0.08 | 0.19 |
| Broad depression | 119 | 1.25 | 0.81 | 1.95 | 0.32 | 0.32 |
| Bipolar disorder | 119 | 1.38 | 0.85 | 2.27 | 0.20 | 0.26 |
| ADHD | 119 | 1.48 | 0.93 | 2.43 | 0.11 | 0.18 |
| ASD | 119 | 1.47 | 0.91 | 2.44 | 0.12 | 0.18 |
| Schizophrenia | 119 | 0.87 | 0.54 | 1.40 | 0.57 | 0.57 |

ADHD: attention deficit hyperactivity disorder; ASD: autism spectrum disorder; CI: confidence interval; FDR: false discovery rate; MDD: major depressive disorder; OR: odds ratio; PGS: polygenic score.

In the South Asian ancestry group, there was also not strong evidence of associations between non-transmitted PGS and emotional disorder (**Table 2**). There was weak evidence of associations between non-transmitted PGS and symptom scores at age 7 (MDD PGS) and age 17 (PGS for anxiety, MDD, broad depression and bipolar disorder), however these results did not withstand FDR correction (**Figure 3b, Table S6**).

#### Neurodevelopmental PGS

There was also not strong evidence of association between non-transmitted neurodevelopmental PGS and any outcome in the European group (**Table 2**, **Figure 3c, Table S5**).

For the South Asian ancestry group, there was weak evidence of association between non-transmitted PGS for ADHD and self-reported symptoms scores at age 17 (SDQ-E and KMH, **Figure 3d**), however, these association did not survive FDR correction (**Table S6**).

### Sensitivity analysis

Sex-stratified analyses did not find strong evidence of sex interaction in the association between any PGS and emotional disorder in the EUR sample (transmitted PGS **Table S7,** non-transmitted PGS **Table S8**). In the SAS sample there was evidence of a sex interaction present for the association between transmitted PGS for MDD and broad depression and emotional disorder, whereby associations were present in males (MDD: OR = 1.8 [1.2,2.7], p_FDR_=0.01; Broad depression: OR=1.71 [1.2,2.5], p_FDR_=0.01), but not females (MDD: OR=0.94 [0.65,1.35], p_FDR_=0.86; Broad depression: OR=0.84 [0.62,1.1], p_FDR_=0.86, **Table S9**). There was not strong evidence of sex differences in the association between any non-transmitted PGS and emotional disorder in the SAS sample (**Tables S10**). There was also not strong evidence of parent (maternal/paternal)-specific genetic nurture in either the European (**Tables S11**) or South Asian (**Tables S12**) ancestry groups.

We investigated evidence for non-random mating in the SAS and EUR ancestry groups to understand whether this may have influenced the results. There was a higher percentage of related parents in the SAS sample (5% third degree [IBD ∼ 12.5%] and 10% fourth degree relatives [IBD ∼ 6.25%]) compared to the EUR sample (<0.1% fourth degree). Correlations between transmitted and non-transmitted PGS, and mother and father PGS are presented in **Figure S1**. Correlations between mother and father PGS and between transmitted and non-transmitted PGS in the SAS sample were higher than in the EUR sample, which could be explained by higher rates of consanguinity or other factors (e.g. assortative mating) in this group and thereby potentially inflating any observed genetic nurture effects.

Analysis of the association between transmitted PGS with emotional disorder in the subset of complete trios, showed a similar pattern of results as in the primary sample (**Tables S13 & S14**), with the exception of an association between transmitted schizophrenia PGS and emotional disorder in the SAS sample (OR=2.4 [1.3,4.5], p_FDR_=0.02), which was not observed in the primary analysis (**Table 1**).

## Discussion

We examined the contribution of psychiatric and neurodevelopmental risk alleles to emotional problems in children and young people, as indexed by transmitted and non-transmitted (i.e. genetic nurture) PGS. In the largest ancestry group (i.e. European ancestry), our results indicate associations between transmitted depression and anxiety risk alleles and youth emotional problems across different measures, informants, ages, and whether defined as continuous or categorical measures. Transmitted neurodevelopmental (particularly ADHD and ASD) risk alleles were also associated with emotional problems. In contrast, we observed little to no association between non-transmitted risk alleles and youth emotional problems indicating a lack of support for genetic nurture.

In keeping with findings from previous twin studies, that emotional problems are heritable with increasing heritability from childhood to adolescence, our results show associations between transmitted psychiatric and neurodevelopmental PGS and emotional problems, particularly during adolescence. The MDD PGS^30^ generally showed the strongest associations, closely followed by PGS for broadly defined depression^31^ and anxiety^29^, with the variance in emotional problem scores explained by these PGS increasing from ages 5 to 17 (0.17-0.61% for anxiety, 0.16-0.97% for MDD, and 0.19-0.91% for broad depression). Bipolar disorder PGS^32^ did not show any robust associations with youth emotional problems. These findings are consistent with studies of youth showing associations between PGS for mood/anxiety disorders and adolescent emotional problems, but not for bipolar disorder PGS^39, 40^. As well as anxiety and depression PGS, we also observed associations of transmitted PGS for ADHD and ASD with youth emotional problems, consistent with previous studies suggesting a neurodevelopmental genetic contribution to some forms of depression^19, 41–43^, (especially early-onset^18^ and treatment resistant^44^) as well as to anxiety symptoms/disorder^45^. It is of note that all cases with a probable emotional disorder in this study can be considered to have “early-onset” emotional problems, since they onset in youth, prior to, or at age 17.

In this study, we found very limited evidence of genetic nurture relating to parental psychiatric/neurodevelopmental genetic liability in the largest (i.e. European) ancestry sample. This suggests that genetic liability to anxiety and depression, as indexed by psychiatric/neurodevelopmental PGS, primarily exerts risk effects on youth depression/anxiety through inherited genetic effects rather than via non-transmitted “nurture” effects. A previous trio-based study, using a different analytical approach, found that parental genetic nurture explained 14% of the variance in parent-reported depression symptoms in offspring at age 8, although not for anxiety symptoms^16^. This previous study examined the total common variant contribution to parent-rated anxiety and depression symptoms at a young age (using the relatedness disequilibrium regression [RDR] approach^46^), whereas our study focused only on the contribution of psychiatric/neurodevelopmental common risk alleles, weighted based on strength of association in previous GWAS. This methodological difference may explain why we observe a much smaller magnitude of variance in offspring mental health explained by non-transmitted MDD PGS (e.g. 0.1% of SDQ-E at age 7), compared to the previous study using the RDR approach (14% of SMFQ at age 8)^16^. Other differences in methodology include the measure of emotional problems used.

Based on our study, we can conclude that non-transmitted common genetic variants that specifically confer risk to psychiatric and neurodevelopmental disorders like depression, anxiety, ADHD, and ASD do not appear to exert strong effects on offspring emotional problems. The non-transmitted PGS approach we used in the present study has previously yielded evidence of genetic nurture effects in the context of childhood educational attainment^12^, and can be considered a complementary approach to other approaches for examining the cross-generational transmission of mental health problems.

At first glance, our findings may seem inconsistent with previous genetic epidemiological findings which have suggested that rearing effects, as well as genetic factors, contribute to emotional disorders. Adoption, children-of-twin and assisted conception design studies observe that environmental, as well as inherited factors, contribute to the cross-generational transmission of anxiety^10^ and depression^11, 47, 48^. However, it is important to recognize that the genetic epidemiological and molecular genetic trio-based designs use different approaches and do not test identical hypotheses.

Genetic epidemiological studies assess *phenotype* resemblance across generations (e.g. parent and offspring emotional problems) to infer genetic and environment contributions, whereas the trio-based design focuses on association of psychiatric/neurodevelopmental risk alleles (transmitted and non-transmitted PGS, rather than parent phenotype) with offspring phenotype. Non-transmitted PGS only capture a proportion of rearing effects: environmental factors associated with parental psychiatric/neurodevelopmental genetic liability (known as passive gene-environment correlation), not all environmental effects. Unmeasured contributions to cross-generational transmission include potential genetic nurture effects resulting from common risk alleles that are not associated with psychiatric and neurodevelopmental conditions in existing GWAS, rare genetic variants, and environmental contributions not influenced by parental genotype (e.g. refugee status, neighborhood adversities, independent life events and other psychosocial stressors independent of parent psychiatric genetic liability). Despite these caveats, our results indicate that caution is warranted in assuming that parents with psychiatric/neurodevelopmental genetic liability confer risk of emotional disorders in their offspring through non-transmitted rearing effects.

Although a strength of the MCS cohort compared to other UK population cohorts includes its ethnic diversity and sampling strategy, there are still relatively few individuals from ancestries other than European. In this study we were able to include families from South Asian ancestry. However, the small sample and the fact that there is a lack of relevant discovery GWAS in South Asian populations, means that our analyses of the South Asian sample were less powered to detect associations than our analyses of European ancestry participants. In addition, genetic samples were imputed using European reference panels, therefore the imputation quality may be lower for the South Asian ancestry individuals compared to European ancestry individuals. Nevertheless, it remains important to highlight this and include these analyses. In this study, we observed higher rates of a probable emotional disorder in the South Asian sample compared to the European sample, and previous evidence from MCS indicates different developmental trajectories of emotional problems in ethnic minority children in the UK compared to White children^49^, indicating that further work on diverse cohorts should be undertaken to investigate contributing factors to emotional problems in these samples.

Our analysis of genetic nurture in the South Asian sample indicated possible weak evidence of associations between non-transmitted psychiatric/neurodevelopmental PGS with emotional problem outcomes, with higher estimated effect sizes than in the European sample, although these associations largely did not withstand FDR correction. Given non-zero correlations between transmitted and non-transmitted PGS and between maternal and paternal PGS, as well as the larger numbers of genetically related individuals within this ancestry group, non-random mating in the South Asian sample may have contributed to our estimation of genetic nurture effects in this study^12^. Whilst the correlation between transmitted and non-transmitted PGS and maternal and paternal PGS could also indicate assortative mating, as the proportion of related parents is higher in the South Asian sample, relatedness is a more likely explanation. However, we cannot rule out the possibility that cross-generational transmission is explained by different mechanisms in different ancestries/cultures. Future work is essential to further understand differences in the intergenerational transmission of emotional problems in diverse samples, both within the UK and globally. A further strength of the MCS cohort is that it is a contemporary sample of people born and growing up in the 21^st^ century, which contrasts to older population cohorts.

Limitations of this study include potential biases due to the available samples. Firstly, study participants did not have DNA collected until age 14, therefore the genotyped sample is likely to be influenced by factors associated with attrition, including neurodevelopmental and psychiatric genetic risk^50–52^. Additionally, since investigation of genetic nurture requires data from complete genotyped trios and genotyping was undertaken when children were aged 14, this limits the sample to offspring with both parents alive/responsive to the study at this time point. Our analysis showed some differences between cohort members belonging to complete trios compared to the whole genotyped sample, including a lower proportion of probable emotional disorder and higher SES, potentially limiting the generalizability of the genetic nurture results to those without complete trio data available. Additionally, as discussed above, analyses in the South Asian ancestry sample are likely underpowered to detect associations between transmitted and non-transmitted PGS and emotional problems, owing to the small sample size available, particularly for complete trios.

## Conclusion

Evidence from this study suggests that transmitted alleles related to depression, anxiety and neurodevelopmental disorders, but not bipolar disorder, are associated with youth emotional problems, with an increasing contribution over the course of childhood and adolescence. The results provide little to no support for genetic nurture effects, suggesting that parental polygenic psychiatric/neurodevelopmental liability does not substantially impact on children’s psychopathology through rearing/nurture effects, although we were unable to conclude whether this is consistent across ancestries.

## Data Availability

MCS survey data can be accessed by bona fide researchers through the UK Data Service (https://doi.org/10.5255/UKDA-Series-2000031).

## Acknowledgements

The authors are grateful to the Centre for Longitudinal Studies (CLS), UCL Social Research Institute, for the use of MCS data and to the UK Data Service for making them available. However, neither CLS nor the UK Data Service bear any responsibility for the analysis or interpretation of these data. This work was supported by the Wolfson Centre for Young People’s Mental Health, established with support from the Wolfson Foundation. We also acknowledge the support of the Supercomputing Wales project, which is part-funded by the European Regional Development Fund (ERDF) via Welsh Government. JM was supported by a NARSAD Young Investigator Grant from the Brain & Behavior Research Foundation (grant no. 27879). CML is part-funded by the NIHR Maudsley Biomedical Research Centre at South London and Maudsley NHS Foundation Trust and King’s College London. The views expressed are those of the author(s) and not necessarily those of the NIHR or the Department of Health and Social Care.

## Author contributions

AS: study design, analysis, interpretation, writing, first author

JM: study design, analysis, interpretation, writing, first author

CAD: study design, analysis, interpretation, editing

LR, MOD, CML: study design, interpretation, editing

AT: study design, interpretation, editing, senior author

