## Supplementary Material for "Estimating the impact of transmitted and non-transmitted psychiatric and neurodevelopmental polygenic scores on youth emotional problems"

### Supplementary Text

#### Genetic quality control

*Millennium Cohort Study (MCS) genetic data*

Saliva samples were collected (using Oragene kits) from MCS cohort members (CMs) and their biological parents, at the age 14 data sweep in 2015–2016; for details of data collection and genotyping, see the published report (1). In brief, a total of 21,432 DNA samples from 21,418 individuals were genotyped using the Infinium global screening arrays-24 v1.0 from Illumina. Genotype calling for 21,368 individuals was performed using Genome Studio v2.0.4. Individuals were excluded if they had a missingness proportion >20% or an estimated heterozygosity deviating from the population mean by 5 standard deviations (N=207 excluded). Samples were imputed using the Michigan Imputation Server; samples were phased with Eagle.v2.4 (2) and imputed to Haplotype Reference Consortium (HRC) release 1.1 (3) using Minimac.v4 (4).

Following imputation, the imputed dosage data were converted to best guess genotypes (binary format) using PLINK-2 (5), applying the following quality control (QC) filters: genotype probability <0.9 per individual, missingness>0.02, minor allele frequency (MAF) <0.01, Hardy-Weinberg equilibrium (HWE) $p<10^{-4}$, and imputation information score (INFO) <0.8. Duplicate position and multiallelic SNPs were excluded. Individuals with excessive missingness (>0.05) were excluded (N=97).

Sex chromosomes were not imputed; sex checks based on provided phenotypic sex were performed using the available genotype data for X and Y chromosomes after basic QC (MAF<0.01, missingness>0.01) and linkage disequilibrium (LD)-pruning (--indep-pairwise 1500 150 0.2). Due to substantial relatedness in the parental generation in the sample, the F statistic was set higher than the default (--check-sex 0.6 0.8). Samples flagged as showing inconsistent phenotypic and chromosomal sex are suggestive of either a sample mix-up during genotyping or an inaccurately recorded phenotype. All such samples were noted and their relationships were inspected, to determine if a resolution could be reached (see below).

Relatedness analyses were performed using the KING software version 2.2.7 (6), to check family relationships by estimating kinship coefficients and inferring identity-by-descent (IBD) segments for all pairwise relationships. For duplicate samples with identical family and person IDs (N=100 pairs), the sample with the higher call rate was kept. For duplicate samples that had different IDs (N=13 pairs), all relationships were inspected and the sample with expected relationships with individuals with the same family ID was kept. All other relationships were inspected, and exclusions were made where samples were mixed up and no resolution could be reached (e.g. 2 CMs with an inferred parent-offspring relationship and evidence of sample mix-up based on sex check or no other relationships in the sample); N=13 excluded. Sample IDs or sex were updated if a sample's identity could be confirmed with certainty (N=60 mixed-up sample IDs were updated and phenotypic sex was updated for 10 confirmed parent samples). Only samples that showed no evidence of sample mix-up and belonged to CMs (based on provided person IDs) or confirmed biological parents of the CMs were retained (N=18,953, including N=8,073 CMs). Mother-father-offspring trios and parent-offspring duo relationships were inferred based on the IBD file to derive sample pedigree information. Mendel analyses in PLINK were used to exclude SNPs with excessive Mendelian errors (>10) and all remaining Mendelian errors were set to missing. Non-transmitted parental alleles were derived for complete trios (N=3,378) using PLINK (--tucc).

Ancestry of the CMs and biological parents was identified using the GenoPred pipeline (7) (available at: https://github.com/opain/GenoPred) in order to derive homogenous ancestry sub-groups using a reference-standardised approach. Through the pipeline, principal components analysis (PCA) was conducted in a reference sample of known ancestry, 1000 Genomes, on independent SNPs (--indep-pairwise 1000 5 0.2). An elastic net model was used to predict the ancestry of the reference sample using the PCs. PCs were then calculated for CMs and biological parents by projecting PCs from the reference sample onto the target sample and applying these PCs to the elastic net model to predict ancestry. By calculating PCs in the target sample in this manner, individual scores are independent of the other members of the sample and are not affected by relatedness within the sample. The elastic net model calculates a probability of each individual belonging to each of the 5 ancestral super-populations (European [EUR], South Asian [SAS], East Asian [EAS], African [AFR] and American [AMR]), and assigns each individual to a group based on their most likely group membership. Individuals with <50% probability of being in their assigned population were excluded. Individuals of EAS, AFR and AMR ancestry were excluded from genetic analyses due to insufficient sample size. QC was performed on CMs from both of the remaining ancestry subgroups (EUR, and SAS) using the GenoPred pipeline. K-means clustering was applied to the sample of CMs to define ancestry clusters; outliers were identified and removed based on their distance from the centroid of their assigned cluster.

There were 68 pairs of CMs, comprised of 133 individual CMs, who were related to each other (MZ, DZ/full sibling, half-sibling/cousin). One CM in each related family was retained, with CMs prioritised on the basis of being in a complete trio and by higher call rate, leading to the exclusion of 67 CMs. Further, the AFR subgroup were excluded from further analysis due to an insufficient sample size for trio analysis (n=33).

The final sample consisted of N=7,142 CMs, which included 3,063 complete parent-offspring trios; see **Table S1** for sample sizes split by ancestry. PCA was re-run on unrelated CMs using the --pca flag in PLINK v1.9 (5), following pruning of the data and removal of regions of the genome with long range LD. PCA was run separately in each ancestry sample (SAS or EUR) to obtain ancestry-specific covariates for analyses.

#### Polygenic score calculation

Allelic information was checked against the HRC reference panel and any SNPs with non-matching alleles were excluded. Palindromic or ambiguous (CT/AG) variants were also excluded. Only common SNPs (MAF>0.01) were kept. This resulted in a sample of 3,976,009 SNPs passing all the above QC.

Discovery summary statistics from the largest available published GWAS were used for 7 phenotypes (see **Table S15** for details). The summary statistics were processed to perform QC filtering, align SNPs against the HRC reference panel and convert summary data to a standardised format, using an R pipeline (available at https://github.com/CardiffMRCPathfinder/summaRygwasqc). None of the GWAS used to generate summary statistics included MCS cohort members.

Polygenic scores (PGS) were calculated for each discovery phenotype in PLINK using the PRS continuous shrinkage (CS) approach (8). This is a polygenic scoring approach that uses all available HapMap-3 (9) SNPs and adjusts the per-SNP effect sizes relative to their GWAS association signals, taking into account LD, using pre-computed LD information provided with PRS-CS, based on the European ancestry subset of the 1000 genomes phase 3 reference sample. PGS were calculated by summing the number of alleles (weighted by the adjusted effect size) across the full set of SNPs for each person. The PRS-CS-auto approach was used, which automatically detects the sparseness of the genetic architecture for each discovery phenotype, based on the discovery summary statistics. The total number of common autosomal SNPs included in each PGS and the weighted mean value of the shrinkage parameter *phi* (weighted by chromosome size) are shown in **Table S16.**

#### Relatedness in ancestry subgroups

Correlations between PGS are presented in **Figure S1**. We see that correlations between $PGS_T$ and $PGS_{NT}$ (top bolded square) as well as correlations between maternal and paternal PGS (bottom bolded square) are more highly correlated in the SAS sample than the EUR sample.

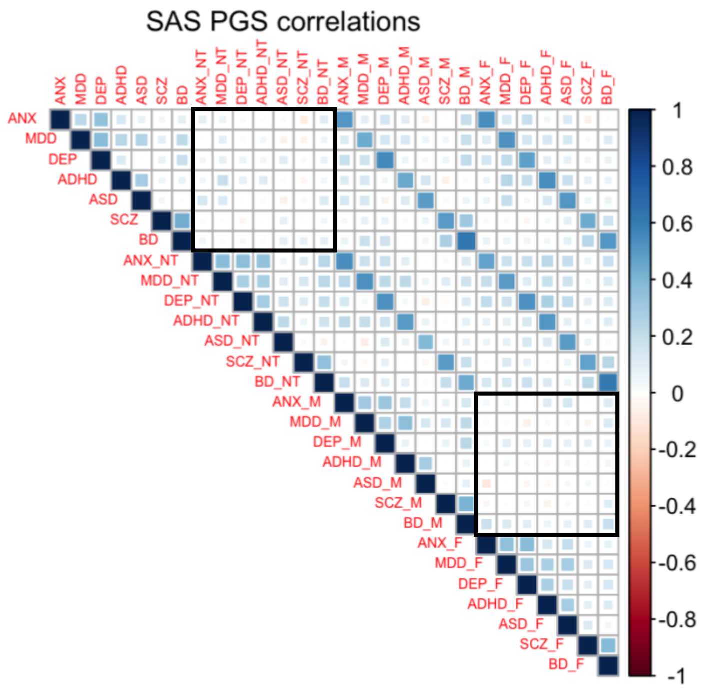

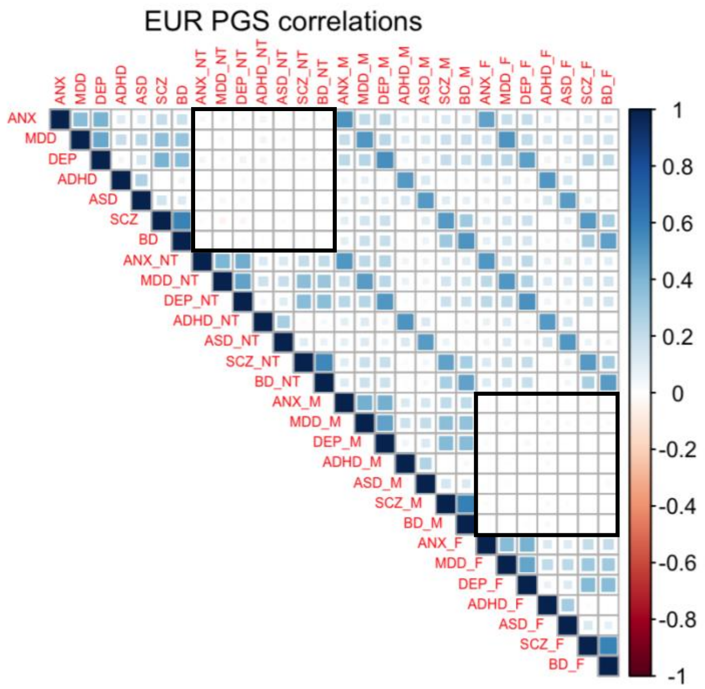

**Figure S1** - *Correlations between polygenic scores (PGS) stratified by ancestry. NT indicates non-transmitted PGS, M indicates maternal PGS, and F indicates paternal PGS. All PGS have been residualised against 10 ancestry-specific principal components. Black bolded squares indicate correlations between transmitted and non-transmitted PGS, and maternal and paternal PGS, used to consider whether relatedness/consanguinity and/or assortative mating influenced SAS results. ASD, autism spectrum disorder; ANX, anxiety; BD, bipolar disorder; DEP, broad depression; MDD, major depressive disorder; SCZ, schizophrenia.*

#### Supplementary Tables

*See document Supplementary_tables.xls for tables.*

Supplementary Table 1 - Comparison of complete trios vs. whole sample.Supplementary Table 2 - Details of missing data.

Supplementary Table 3 - EUR subgroup associations between transmitted PGS and childhood emotional problem symptom scores.

Supplementary Table 4 - SAS subgroup associations between transmitted PGS and childhood emotional problem symptom scores.

Supplementary Table 5 - EUR subgroup associations between non-transmitted PGS and childhood emotional problem symptom scores.

Supplementary Table 6 - SAS subgroup associations between non-transmitted PGS and childhood emotional problem symptom scores.

Supplementary Table 7 - Sex-stratified EUR subgroup associations between transmitted PGS and emotional disorder in youth.

Supplementary Table 8 - Sex-stratified EUR subgroup associations between non-transmitted PGS with emotional disorder in youth.

Supplementary Table 9 - Sex-stratified SAS subgroup associations between transmitted PGS and emotional disorder in youth.

Supplementary Table 10 - Sex-stratified SAS subgroup associations between non-transmitted PGS and emotional disorder in youth.

Supplementary Table 11 - EUR subgroup parent-specific associations between non-transmitted PGS and emotional disorder in youth.

Supplementary Table 12 - SAS subgroup parent-specific associations between non-transmitted PGS and emotional disorder in youth.

Supplementary Table 13 - Complete trios only EUR subgroup associations between transmitted PGS and emotional disorder in youth.

Supplementary Table 14 - Complete trios only SAS subgroup associations between transmitted PGS and emotional disorder in youth.

Supplementary Table 15 - Summary statistics from the following published GWAS used to generate PGS in the MCS sample.

Supplementary Table 16 - Details of the number of autosomal SNPs included and the weighted mean value of the shrinkage parameter (phi) used for the generation of each PGS.
